# Containing Covid-19 outbreaks with spatially targeted short-term lockdowns and mass-testing

**DOI:** 10.1101/2020.05.05.20092221

**Authors:** Justin Alsing, Naïri Usher, Philip JD Crowley

## Abstract

We assess the efficacy of spatially targeted lockdown or mass-testing and case-isolation in individual communities, as a complement to contact-tracing and social-distancing, for containing SARS-CoV-2 outbreaks. Using the UK as a case study, we construct a stochastic branching process model for the virus transmission, embedded on a network interaction model encoding mobility patterns in the UK. The network model is based on commuter data from the 2011 census, a catchment area model for schools, and a phenomenological model for mobility and interactions outside of work, school, and the home. We show that for outbreak scenarios where contact-tracing and moderate social distancing alone provide suppression but do not contain the spread, targeted lockdowns or mass-testing interventions at the level of individual communities (with just a few thousand inhabitants) can be effective at containing outbreaks. For spatially targeted mass-testing, a moderate increase in testing capacity would be required (typically < 40000 additional tests per day), while for local lockdowns we find that only a small fraction (typically < 0.1%) of the population needs to be locked down at any one time (assuming that one third of transmission occurs in the home, at work or school, and out in the wider community respectively). The efficacy of spatially targeted interventions is contingent on an appreciable fraction of transmission events occurring within (relative to across) communities. Confirming the efficacy of community-level interventions therefore calls for detailed investigation of spatial transmission patterns for SARS-CoV-2, accounting for sub-community-scale transmission dynamics, and changes in mobility patterns due to the presence of other containment measures (such as social distancing and travel restrictions).

## 1 Introduction

The novel coronavirus SARS-CoV-2 was first detected in Wuhan in late 2019^1,2^ with human transmission observed soon after^3,4^ and was found to cause severe disease in some cases. Despite containment efforts, the virus spread across the globe, resulting in a global pandemic with over 2.5 million confirmed cases and 170,000 deaths worldwide as of 23 April 2020. In order to contain or mitigate the spread of the virus, countries have deployed various combinations of measures, including tracing and isolation of contacts, social-distancing, and in many cases stringent lockdowns^5,6^. The heterogeneity of the implemented strategies across countries reflects the fact that the optimal approach is unknown, and likely context dependent.

Contact-tracing in particular has gained significant attention, having played a notable role in containing the outbreak in South Korea and Hong Kong (and elsewhere), and having a strong recent history including with severe acute respiratory syndrome (SARS)^7^, Middle East respiratory syndrome (MERS)^8,9^, Ebola^10,11^, and others^12,13^. This has led to many countries developing digital contact tracing apps to enable automated reporting of symptoms and tracing of contacts via smartphones^14^.

The efficacy of contact-tracing as a containment strategy is dependent of four key factors: the proportion of transmission that occurs before symptom onset (and from asymptomatic cases)^15–17^, the basic reproduction number^16,17^ (*R*_0_), the delay from symptom onset to isolation^14,16,17^, and the fraction of contacts that are successfully traced and quarantined or isolated^14,16–18^. SARS-CoV-2 presents a particular challenge due to its relatively high reproduction number (*R*_0_ ≃ 3 − 4^5,19,20^), significant pre-symptomatic^21,22^ and possibly asymptomatic^23,24^ transmission. This puts onerous requirements on any digital (or other) contact-tracing effort to trace a high fraction of contacts, and isolate them promptly. It is estimated that for the SARS-CoV-2 pathogen, contact-tracing alone is only able to achieve containment with around 80% of contacts traced^14,17^, rapid isolation and/or quarantining of large numbers of traced contacts who do not yet show any symptoms^14,18^, and a modest number of imported cases^17^. Even for countries with high smartphone ownership, this is likely unrealistic for digital contact tracing alone; in the UK for example, 80% of adults own a smartphone^25^, which naively puts an upper limit of 64%^1^ on the number of contacts traced through digital contact tracing (even if app adoption was mandatory). Digital contact tracing is hence unlikely to be enough to achieve containment on its own, and additional measures will be necessary, including (some combination of) supplementary manual contact tracing, social distancing measures and travel restrictions, isolation of individual households, schools, or workplaces, mass testing^26^, and possibly community-level interventions^27^, etc.

In this study we investigate the efficacy of spatially-targeted testing and case isolation at the level of small communities (of a few thousand inhabitants), as a complement to contact tracing for SARS-CoV-2 containment. Testing and lockdown interventions are implemented in dynamically identified infection hotspots when the number of local cases is identified to pass a threshold. We use a stochastic branching process transmission model (calibrated to SARS-CoV-2), embedded on a network interaction model describing mobility patterns between communities in the UK^2^, based on 2011 census data for commuter flows, a catchment model for schools, and a phenomenological model for mobility and interaction outside of school or the workplace. This allows us to simulate the impact of community-level intervention measures on containing outbreaks.

For both intervention measures explored (spatially targeted lockdowns or mass-testing), we track the effective reproduction number in each community using the (partially) observed daily case-numbers as the outbreak grows. Once the effective reproduction number is observed to pass some threshold in a given community, an intervention measure is triggered. Local lockdown interventions are simulated as fixed-term (two week) lockdowns on individual communities, during which transmission is suppressed in-line with observations of SARS-CoV-2 transmission under lockdown across Europe^5^. For targeted mass-testing interventions, we simulate testing an entire community over three days and subsequently isolating positive cases, whilst accounting for a realistic false negative rate^28,29^.

Using England and Wales as a case study, our anaylsis indicates that for scenarios where contact-tracing and modest social distancing alone achieves some suppression but fails to contain the spread, imposing additional targeted lockdowns or mass-testing at the community-level can be effective at containing outbreaks, with a small fraction (typically <0.1%) of the population locked down at any one time, or a moderate increase (typically < 40000/day) in testing capacity.

## 2 Model

We implemented a branching process model for the virus transmission (following closely Hellewell et. al. 2020^17^). The branching process was embedded on a network interaction model encoding mobility patterns in the UK based on commuter data from the 2011 census, a catchment area model for children traveling to school (using the locations of UK schools), and a phenomenological (gravity law) model describing peoples movements and interactions outside of school, work or the home.

### 2.1 Transmission model structure

A number of initial cases (parent nodes) are introduced into the country at the beginning of the outbreak (randomly distributed spatially). Each case gets an incubation period, drawn from an incubation-period distribution. We assume that some time after symptom onset, infectees will self-isolate with an onset-to-isolation delay drawn from an isolation-delay distribution. Each case then generates a number of potential secondary infections (child nodes) drawn from a negative binomial with mean equal to the basic reproduction number, and heterogeneity in the number of generated infections (ensuring the possibility of super-spreader events^30^). The infection times of each potential secondary infection are drawn from a generation time distribution, conditioned on the symptom onset time of the parent. Secondary cases are then only accepted if their infection times occurred before their parent node self-isolated; isolation is assumed to be completely effective at preventing further transmission. The communities (ie., geographical locations) of each secondary case is drawn from a network interaction model, given the community and occupation of the parent node (see ‘Network interaction model’ below). Finally, cases are terminated (recovery or death) some time after symptom onset, drawn from the termination time distribution. The assumed incubation period, isolation-delay, offspring number, generation time and termination time distributions are calibrated to observations for the SARS-CoV-2 pathogen and detailed below (see ‘SARS-CoV-2 parameter assumptions’).

### 2.2 Contact tracing

We assume that a fraction *ρ* of cases are successfully contact traced, and traced cases are assumed to self-isolate immediately on becoming symptomatic, as opposed to after some delay. Note we do not currently include quarantine of pre-symptomatic cases.

### 2.3 Local lockdown and mass-testing measures

In addition to contact tracing and isolation, we allowed for an additional intervention in the form of either locking down or mass-testing and isolating cases in individual communities (the definition of community is discussed under ‘Network interaction model’ below).

As the outbreak spreads, we assume that some fraction of cases are confirmed (via testing) and reported at the time when infectees isolate (either spontaneously after some delay, or immediately on symptom onset if they have been contact traced as described above). The live confirmed case data for each community is then used to define an intervention threshold, where either a fixed-term lockdown or mass-testing and isolation is triggered when a community passes this threshold.

Choosing the optimal threshold for imposing local interventions given the case data in each community, knowledge of inter-community mixing and possibly even reconstructed movements of infectious individuals and their contacts, is a complex decision task. In this case study we show that even with a simple (sub-optimal) intervention threshold, local lockdowns (or mass-testing) can act as an effective compliment to contact tracing for outbreak containment.

We implemented a simple intervention threshold, as follows. During simulations, at the end of each full simulated day we construct an estimator^3^ (and associated uncertainty) for the effective reproduction number *R*_eff_ for each community given the past week of case reports. If the estimated *R*_eff_ > 1 with confidence exceeding 84% for any community, then a local intervention is triggered. While this simple threshold criteria is sufficient for our proof-of-concept study, we stress that in reality a sophisticated inference engine for monitoring the infection surface should be used for triggering local interventions.

For local lockdown interventions, we impose a fixed-term two-week lockdown on individual communities. During lockdowns we assume the effective reproduction number for members of that community is reduced by 80%, consistent with recent measurements of the impact of lockdown measures in Europe on SARS-CoV-2 transmission^5^.

For local mass-testing and isolation interventions, we assume that the entire community (of a few thousand inhabitants) can be tested over the course of three days, with positive cases subsequently isolated. For the three-day testing period, the community is put under lockdown during which we assume transmission is reduced by 80%. We assume a false negative rate of 20% for testing, so only 80% of active infections are successfully identified and isolated during a mass-testing intervention.

### 2.4 Network interaction model

England and Wales are divided into communities using the ‘Middle Layer Super Output Area’ (MSOA) system (a geospatial statistical unit used for reporting small area statistics). There are 7100 MSOAs in England and Wales^4^, each with a population of between five and sixteen thousand people.

The population is divided into three occupation types — people who work (W), school children (S), and people who neither go to work or school (O) — and their relative numbers in each MSOA is fixed by the 2011 census data. We define the interaction strength *I_i j_* as the probability that someone from community *i* will infect someone from community *j* in a given transmission event, and the interaction matrices are assumed to be heterogeneous with respect to both infector type (worker, school child or other), and the location of the transmission event (at work, at school, at home, or in the community).

We assume that one third of transmission occurs in the home (h), one third happens at school (s) or in the workplace (w), and one third out in the community (c). This is consistent with mixing patterns reported in social mixing studies^31^ (and assumed elsewhere^26^). The interaction model for inter-community transmission in each of these contexts is then constructed as follows.

#### Workplace transmission

Each parent node in the branching process is assigned an occupation-type based on the relative fractions of W, S and O in their home community (from the 2011 census data). If a parent node (from community *i)* is a worker (W), their workplace community *k* is drawn randomly using the 2011 commuter data (ie., the number of people commuting from their home community to each MSOA). Their work-time interaction matrix is then determined by the relative populations of commuters in their workplace community *k*, from each other community *j*, ie.,

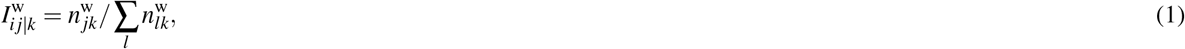

where 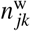 is the number of people commuting from community *j* to community *k* for work (taken from the 2011 census data). We define workplace transmission to only occur between workers.

#### School transmission

If a parent node is a school child (S), their school community *k* is drawn assuming a school catchment model as follows. England and Wales are first further divided into LSOAs^5^ — smaller communities with an average population of around fifteen hundred. Schools are placed into LSOAs based on the known locations of schools in England, and placed randomly (tracing the population density) in Wales^6^. School children in each LSOA are then assumed to attend one of the nearest six schools with equal probability^7^; this defines an expected flow of school children between LSOAs for school attendance. This LSOA level flow is then downgraded onto the MSOA communities to obtain the expected number of school children traveling between each MSOA for school. The school-time interaction matrix for a child from community *i* who attends school in community *k*, is then given by,

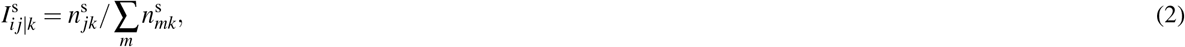

where 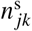 is the number of people commuting from community *j* to community *k* for school based on the catchment model, and we assume that school children only interact with (and are well mixed with) other school children during school-time.

#### Home transmission

For transmission that occurs in the home, we assume that all interactions are with members of the home community (by definition),

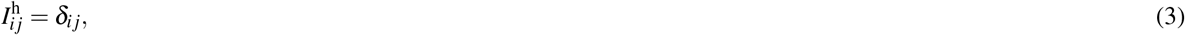

where *δ_ij_* is the knonecker delta.

#### Community transmission

For transmission that occurs out in the community, we assume that outside of work, school or home time people move according to a gravity law model (a class phenomenological models commonly used to describe human flows based on populations and distances between population centres, and widely applied to study the spatial patterns of disease spread^32–34^). Here, we assume the probability of a node from community *i* finding themselves in community *j* is be given by,

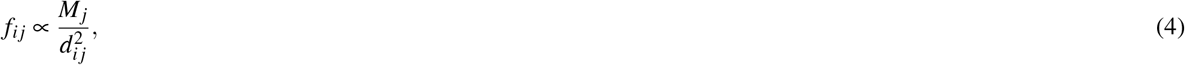

where 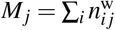 is the working commuter population of node *j*, *d_ij_* the distance between communities *i* and *j*, and the gravity law is normalized such that 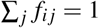. We are assuming that outside of the home, workplace or school, people congregate proportional to the density of commercial activity, for which we take the working (commuter) occupancy as a proxy. This ensures that people mix well in dense urban centers during their leisure time. The inverse-square dependence on distance provides a heavy-tailed distribution of transmission distances, ensuring the possibility of long-range transmission (discussed further below).

The interaction matrix associated with community transmission is hence determined by the expected occupancy of people across communities according to the gravity law:

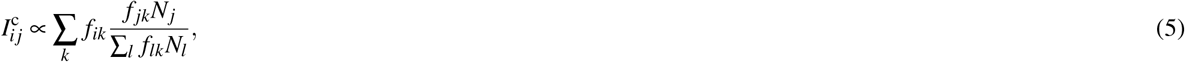

where *N_j_* is the population of community *j*, and the community interaction term is normalized such that 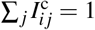.

#### Combined interaction model

The communities of the offspring of a parent node from community *i* are drawn as follows. The occupation of the parent node is chosen according the relative fractions of workers, school attendees and other in their home community. If they are a worker or school attendee, their school or workplace location *k* is chosen according to the catchment model or commuter data accordingly. Then, the locations *j* of their offspring are chosen with probabilities 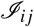 according to (for workers, school children and other respectively):

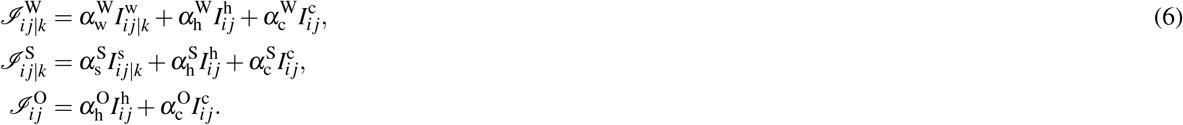

The relative fractions *α* of transmission that occur at home, at school or in the workplace, and in the community for each occupation type are fixed such that for the population as a whole, the overall amount of transmission in the three contexts is one third each^26,31^. The result is that for workers and school children, approximately two-thirds of their transmission occurs at school or in the workplace and one sixth at home or in the community respectively. For those who neither work or attend school, half of their transmission is assumed to occur in the home and in the community respectively.

#### Discussion and caveats

With the assumptions outlined above, for England and Wales as a whole the network model assumes that typically between 30 and 40% of transmission events are intra-community, with dense urban communities exhibiting the lowest levels of intra-community transmission (ie., those communities are more mixed^8^) and rural communities the highest. For transmission that occurs in the community and in the workplace, the intra-community transmission constitutes ≲ 10 − 20%, while in schools intra-community transmission accounts for ~20-40% of the total. This means that the community and workplace interaction terms describe high levels of mixing, school interactions somewhat less so, and home interactions are entirely intra-community.

The gravity model for community interactions (and to a lesser extent, the commuter data) ensures that the distribution of transmission distances has a heavy-tail leading to plausible levels of long-range transmission; 5% of transmission events occur over 100km, 2% over 200km and 1% over 250km.

It is important to assess the sensitivity of the results to the network model assumptions. The efficacy of spatially targeted interventions should be principally driven by the spatial clustering of transmission events, with high levels of intra-community transmission expected to be effectively suppressed by targeted local interventions, and high levels of intercommunity transmission being more challenging to contain (or, demanding that any interventions are made over larger spatial regions).

We do not currently attempt to model heterogeneity in transmission intensity with respect to age^26,35^, or clustering (and saturation) at the sub-community level (ie., individual households, workplaces, or schools^26^). While these effects are undoubtedly important for the details of the epidemic dynamics, we stress that the key macroscopic parameter for estimating the efficacy of community-level interventions is level of intra- versus inter-community transmission. Furthermore, there is still considerable uncertainty in the age heterogeneity for SARS-CoV-2 transmission.

We have estimated mobility patterns assuming that people are going to work and school as usual and are free to move as they please during their leisure time. In practise, with some level of social distancing measures and travel restrictions in place, mobility patterns will differ from those under normal circumstances. This will likely increase the relative fraction of intra-community transmission, making spatially targeted interventions more effective.

Clearly, confirming the efficacy of community-level interventions with confidence will require a detailed investigation of the spatial dynamics of transmission inside and between communities, taking into account as many factors as possible. See ‘Discussion’ for a description of ongoing work.

### 2.5 SARS-CoV-2 parameter assumptions

We assume a Weibull distribution for the incubation period, with a mean of 5.8 and standard deviation of 2.6 days^36^.

The number of offspring for each parent node is drawn from a negative binomial distribution, with mean equal to the basic reproduction number *R*_0_ and dispersion *δ* = 0.16 (calibrated to data for SARS^30^). The reproduction number is expected to be context (country) dependent^5^, and will depend on any social distancing or other measures than are in place to suppress transmission, as well as the amount of herd immunity already attained after an initial outbreak. For our baseline case we take *R*_0_ = 2.5, which is in the expected range for SARS-CoV-2 in the UK with modest social distancing measures in place^5^.

For the onset-to-isolation delays, we assume a Weibull distribution with mean 3.43 and standard deviation 2.4 days, calibrated to the late stages of the 2003 SARS epidemic in Singapore^37^. Expectations for the typical delay between initial onset and developing sufficient symptoms to seek medical care (and testing) or self-isolate, vary considerably. Ferguson et al 2020 suggest that two thirds of cases could feasibly self-isolate within a day of initial symptom onset if required by policy^26^, while data from South Korea, Singapore and Hong Kong indicate that the mean delay from initial onset to the first medical consultation there was 2.1 days (with a standard deviation of 1.65 days)^26^. However, uncertainties in the number of mild cases (who may take longer to self-isolate) persist, and the onset-to-isolation distribution remains hard to predict across varying contexts.

The generation time distribution is taken to be skew normal^17^, centered on the symptom onset time with skewness of 1.95 and standard deviation of 2. This results in 15% of transmission occurring in the pre-symptomatic phase (in the absence of self- or other- isolation measures), infectiousness beginning typically two days before symptom onset, and lasting for around a week after symptom onset. In the presence of self-isolation, the pre-symptomatic transmission fraction increases to 20%, and with both self-isolation and contact-tracing^9^ increases to 70%. This is broadly consistent with estimates from data^38^, although some laboratory montiroing of viral shedding suggests the fraction of pre-symptomatic transmission could be even higher^39^.

Combining the assumed incubation period and generation time distributions leads to serial intervals with mean 7.1 and standard deviation 2.9 days (in the absence of self-isolation). In the presence of self-isolation the mean serial interval reduces to 6.4 days (standard deviation 2.8). This is in line with observed serial intervals from confirmed case-pairs^40,41^ (although some studies suggest shorter serial intervals^14,42,43^).

The case termination time distribution is taken to be a Gamma distribution with mean 24.7 and standard deviation 8.6 days, taken as a fit to the onset-to-recovery times for SARS-CoV-2^44^ (we neglect the correction due to shorter termination times for cases that end in death^44^).

## 3 Scenarios

As a default scenario, we seed outbreaks with 20 initial infections distributed according to population density. We assumed an *R*_0_ = 2.5 which may be expected for SARS-CoV-2 in the UK with modest social distancing measures in place, and that 64% of cases are successfully contact traced and isolated on symptom onset. We note that this represents a theoretical upper limit for digital contact tracing, assuming that 80% of people in the UK own a smartphone, and app adoption was made compulsory (leading to *ρ* = 0.8^2^ = 0.64 if all contacts who own a smartphone were successfully traced).

We run 1000 simulations for three intervention schemes: (a) baseline interventions of contact-tracing and modest social distancing alone; (b) baseline interventions in conjunction with targeted mass-testing and isolation of cases; and (c) baseline interventions in conjunction with targeted lockdowns. The branching process was terminated when the outbreak was either contained (ie., all active cases were successfully isolated) within eight months, the total number of active cases exceeded ten thousand before eight months (for which we assumed the outbreak was out of control), or the branching tree was complete up to eight months but the outbreak ongoing.

## 4 Results

Fig. 1 summarizes the results for the baseline interventions (contact-tracing and modest social-distancing alone), and with the addition of spatially targeted mass-testing and case-isolation.

**Figure 1.**
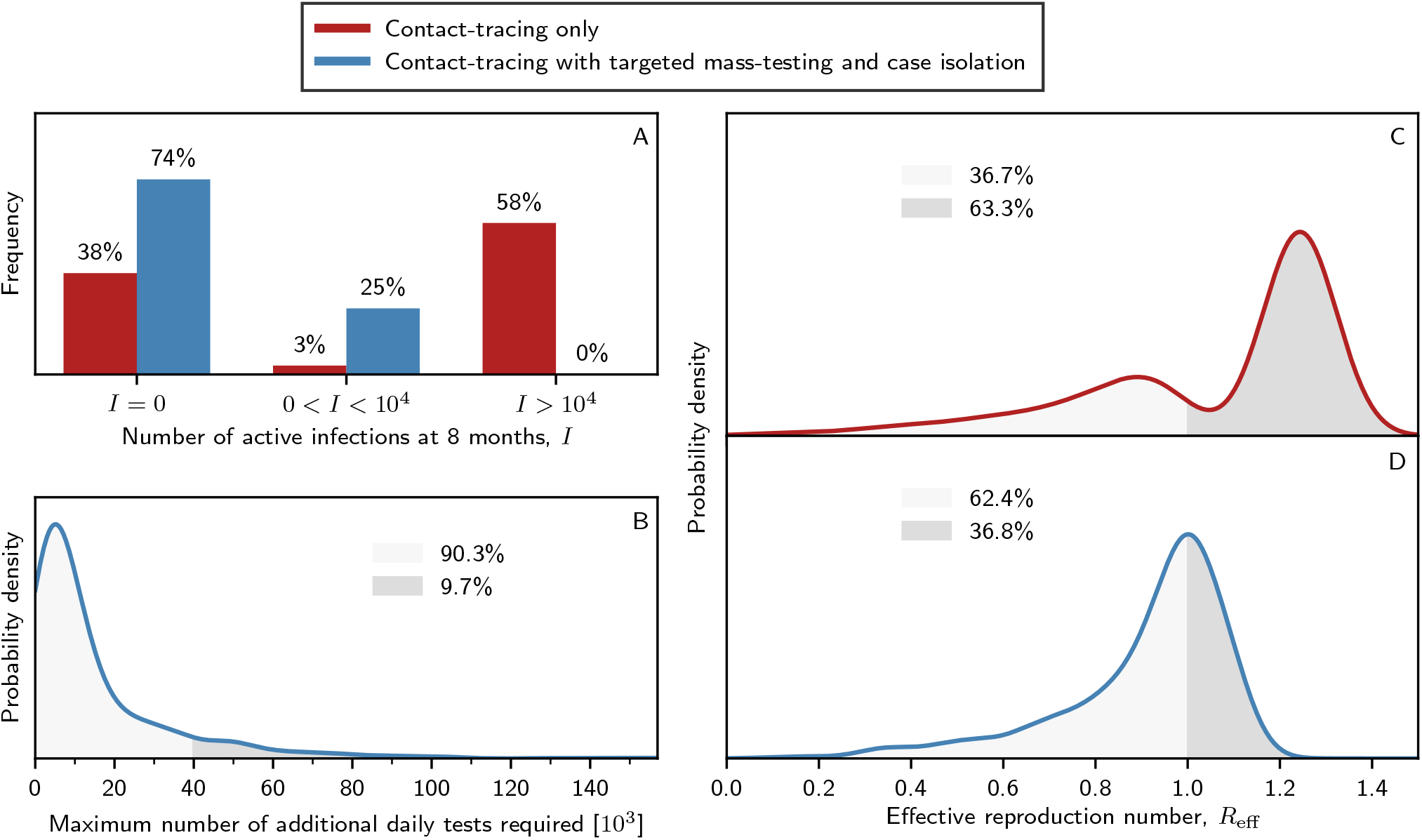
Summary statistics of simulated outbreaks for two cases: contact-tracing and modest social distancing only (baseline), and with the addition of targeted mass-testing and case-isolation. 1000 outbreaks are simulated for each of the two cases. Each simulation is terminated for one of three stopping conditions. (A) The proportion of simulated outbreaks that are terminated for each of the three stopping conditions: the outbreak is contained within 8 months (i.e., zero active infections at 8 months), the outbreak is uncontrolled within 8 months (i.e., the number of active infections at 8 months exceeds the ten thousand case limit), and the outbreak is neither contained nor uncontrolled within 8 months (i.e., the number of active infections at 8 months is less than ten thousand, but the outbreak is ongoing). (B) The maximum number of additional daily tests required to implement the targeted testing protocol. (C) Effective reproductive number of outbreaks for the baseline intervention scenario. (D) Effective reproductive number of outbreaks when targeted testing is used in conjunction with the baseline interventions.

In the baseline case, we find that 38% of the 1000 simulations are contained within eight months, while 58% of simulated outbreaks exceeded the ten thousand case-limit. With the addition of targeted mass-testing, 74% of outbreaks were contained and none of the simulated outbreaks reached the ten thousand case-limit by eight months.

For each simulation, we calculated the effective reproduction number, *R*_eff_, as the average reproduction number per infection. In the baseline case, we find that 63.3% of outbreaks result in *R*_eff_ > 1 (resulting in exponentially growing outbreaks), while with the addition of targeted mass-testing this fraction decreases to 36.8%. We re-iterate that even for cases with *R*_eff_ > 1, with targeted mass-testing interventions none of those outbreaks reach the ten thousand case limit within eight months.

The number of additional daily tests required to implement the targeted mass-testing strategy are shown in Fig. 1 (bottom-left). In 58.8% of simulated outbreaks, the number of additional tests required is less than ten thousand (per day). In 90.3% of simulations, the required additional testing capacity is less than forty thousand per day.

Fig. 2 illustrates typical outbreak scenarios with and without targeted mass-testing (the cases shown are chosen with *R*_eff_ close to the respective modal values). In the baseline case, the most commonly occurring outcome is an uncontrolled outbreak (the simulation terminates when the number of active infections surpasses ten thousand). For the case with targeted mass-testing, the most commonly occurring outcome is a contained outbreak (the simulation terminates when there are no active infections).

**Figure 2.**
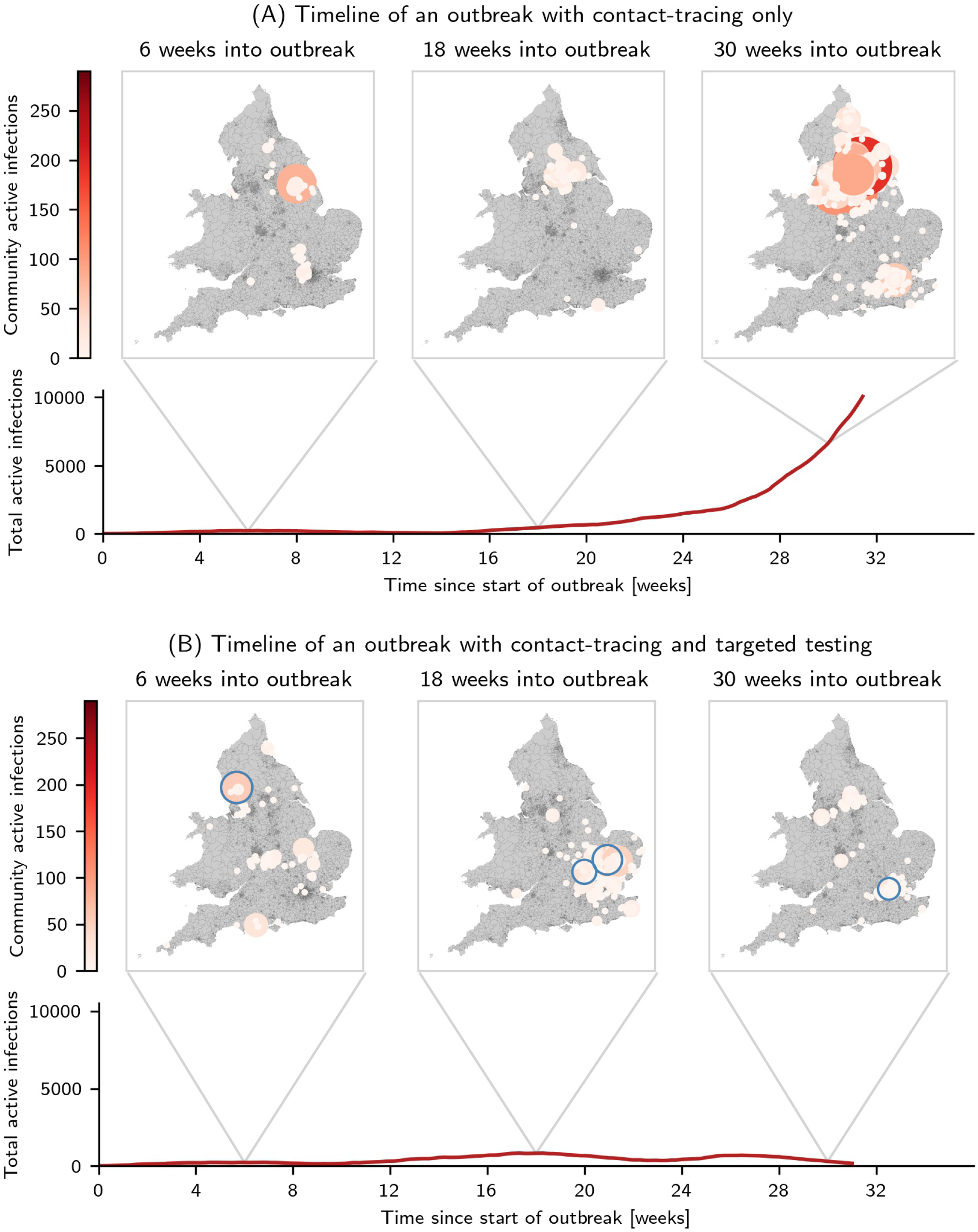
An example of the most common simulation outcome for each of the two cases. For each of the two examples, we show the time series of the total number of active infections in England and Wales. Above the timeseries, we show snapshot images of a map of England and Wales at 6, 18 and 30 weeks into the outbreak. Each coloured circle on the maps shows the number of active infections within a single community. The communities that are currently undergoing the targeted testing protocol as circled in blue. (A) An example simulation from the contact-tracing only case, where the simulation terminates when the total active infections cross ten thousand. (B) An example simulation from the baseline (contact-tracing and social-distancing only) case, where the simulation terminates when the total active infections cross ten thousand.

Fig. 3 shows the summary statistics for the baseline case alongside the targeted local lockdown strategy. As one would expect, we find that the targeted lockdown protocol leads to strong outbreak suppression: 87% of outbreaks are contained (compared with the 74% seen with targeted testing scheme), and 38% with the baseline interventions. The additional suppression comes at the cost of a larger number of people in lockdown at any one time. Nevertheless, in 86% of outbreaks the maximum number of people in lockdown at any one time is less than one hundred thousand, and even in the far tails of the distribution the maximum number of people locked down is less than a few hundred thousand (corresponding to < 1% of the population of England and Wales).

**Figure 3.**
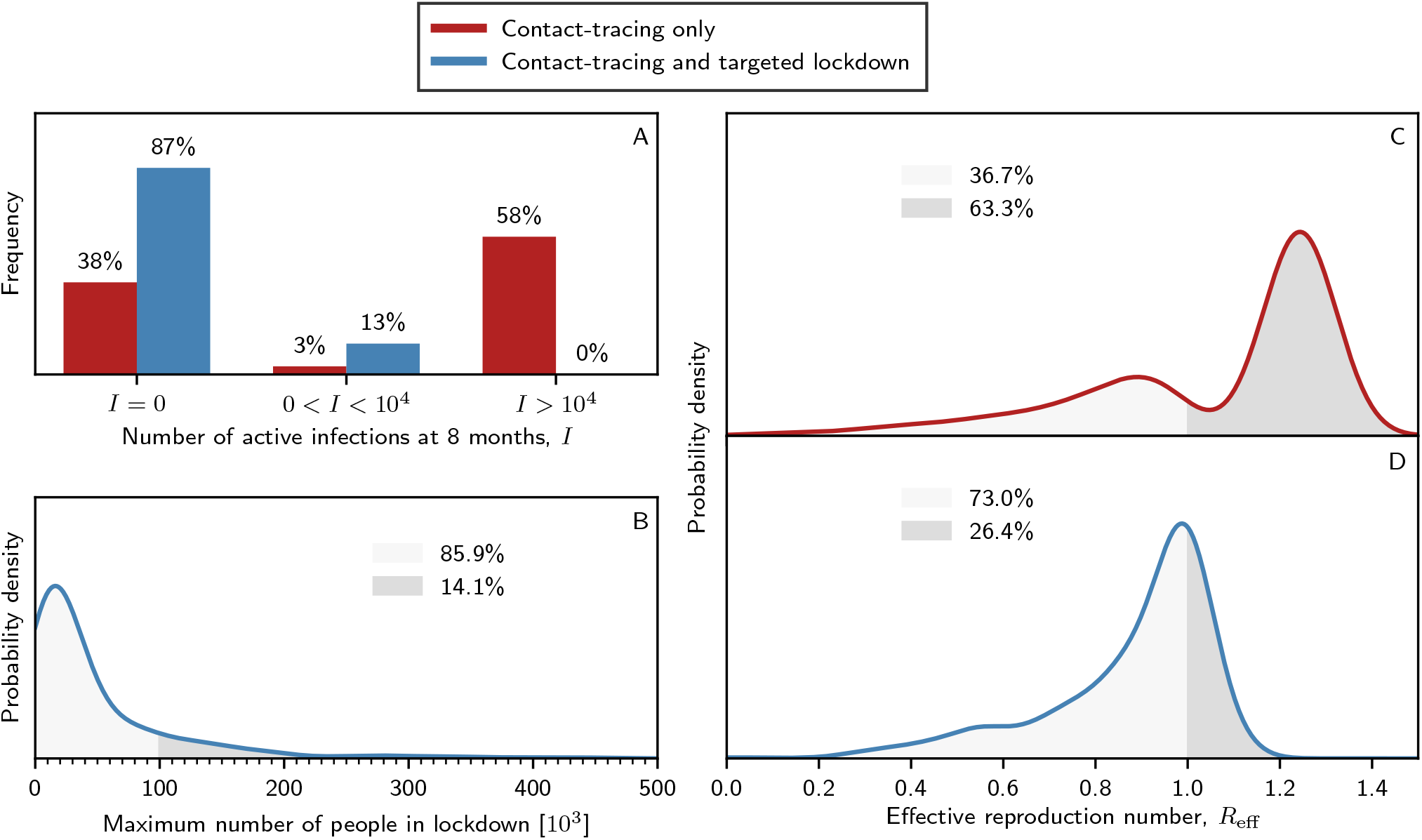
Summary statistics of simulated outbreaks for two cases: contact-tracing and modest social distancing only (baseline), and with the addition of targeted two-week lockdowns at the communuty-level. 1000 outbreaks are simulated for each of the two cases. (A) The proportion of simulated outbreaks that are terminated for each of the three stopping conditions: the outbreak is contained within 8 months (i.e. zero active infections at 8 months), the outbreak is uncontrolled within 8 months (i.e. active infections at 8 months exceeds the ten thousand case limit), and the outbreak is neither contained nor uncontrolled within 8 months (i.e., fewer than ten thousand active infections at eight months but the outbreak is ongoing). (B) The maximum number of people who are in lockdown at any one time in the targeted two-week lockdown protocol. (C) Effective reproductive number of outbreaks for the baseline case. (D) Effective reproductive number of outbreaks when targeted two-week lockdown is used in conjunction with the baseline interventions.

## 5 Discussion

We have developed a model of disease transmission incorporating population density and commuter flow patterns derived from UK census data. Within this framework, we have shown that in situations where an appreciable fraction of transmission occurs within versus across communities, targeted interventions at the community-level (via either lockdowns or mass-testing and isolation) can be effective at containing SARS-CoV-2 outbreaks.

Confirming the efficacy of spatially targeted interventions with confidence requires exploring a number of avenues. First, given the degree of variation in the reported SARS-CoV-2 parameters (Section 2.5), further work is necessary to establish the insensitivity of this conclusion to variation in the parameters. Second, the spatial dynamics of transmission inside and between communities must be carefully interrogated in the context of age and context heterogeneous transmission, sub-community scale effects such as household, school or workplace saturation (as the majority of members become infected), and as mobility patterns change under the presence of other containment measures (including various social distancing and travel restriction scenarios). Finally, while the commuter data and school catchment model should provide a reasonable (data-driven) picture of inter-community interaction patterns at school and in the workplace, our model currently relies on a phenomenological (gravity) model for interaction patterns outside of the home, workplace or school. This could be improved with a data-driven model for mobility patterns outside of those contexts, using eg., social network data^45^.

In addition to understanding the spatial transmission dynamics in detail, the definition of communities and thresholds for triggering local interventions merit exploration. In this proof-of-concept we used simple (and a priori fixed) community definitions and a somewhat crude intervention threshold. Optimal community definitions could involve generating community boundaries with the goal in mind, ie., such that the intra-community transmission is sufficiently high that local interventions should be effective. Community boundaries also need not be static, and could be updated as mobility patterns change under changing social distancing (and other) measures. Intervention thresholds too should take into account the ‘nowcast’ infection surface, and the interventions themselves (eg., which communities are targeted at a given time) may be a function of their inter-connectedness due to mobility patterns.

Ethical issues surrounding spatially-targeted intervention measures merit very careful consideration. It is clearly important to ensure that any containment scheme is both as effective as possible, while avoiding excessive (unfair) burden on any particular group of people. Spatial-targeted interventions are particularly sensitive, because both transmission dynamics and impact (in terms of hospitalizations and deaths) may be expected to correlate with socio-economic factors. While heterogeneous burden sharing resulting from spatially-targeted interventions may be (partially) compensated by appropriate re-allocation of resources, we urge that factoring spatial-targeting into policy decisions deserves considerable care.

### Code and ongoing work

Code is available on request and will be made public on github imminently. We note that the code has greater flexibility than the demonstrations presented in this paper, with full control over asymptomatic cases, and details of the contact-tracing assumptions including both quarantine and isolation efficacy and delays, etc.

We are extending the transmission model to include age and context heterogeneity, with transmission events being underpinned by age and context dependent contact distributions (using the BBC Pandemic dataset^46^), household distributions taken from the 2011 census data, and including the effects of household, school and workplace saturation. The interplay between these sub-community-scale factors and the mobility model should constrain the relative fraction of intra-community transmission with greater confidence, allowing for stronger statements on the efficacy of community-level intervention measures. We are also exploring the use of social network data^45^ for improving the interaction model (in particular, mobility patterns outside of work, school and household contexts).

Finally, we are also looking at how the spatially targeted interventions affect the demand on regional health services. Since the targeted interventions by definition limit outbreaks only in the areas in which outbreaks are most intense, the benefit of the strategy is best measured by the outcomes at a regional level.

## Data Availability

The code associated with the article is available on GitHub

## 6 Acknowledgements

We thank Luke Yeo for helpful discussions throughout, and Anastasios Noulas and Till Hoffmann for a helpful discussion regarding mobility networks.

## Disclaimer

*We stress that this is a working paper where results are preliminary and subject to change. In particular, we note that the efficacy of spatially targeted interventions are sensitive to the relative proportions of intra- versus inter-community transmission (for a given definition of community boundaries), which in turn is sensitive to the assumptions about the transmission dynamics across different contexts. Whilst the assumptions made here about transmission across contexts are motivated, we are currently updating our model to make the estimated inter- and intra- community transmission rates as robust as possible, as well as running a comprehensive suite of sensitivity tests and different outbreak scenarios*.

## Footnotes

1 For a transmission event to be successfully traced, both infector and infectee must be traceable. Therefore, with *ρ*% app adoption in the population, an upper limit of *ρ*^2^% cases could be traceable through digital contact tracing alone.

2 We split the UK into ‘Middle Super Output Areas’ (MSOAs) – communities with between five and sixteen thousand inhabitants.

3 We construct an estimator for the recent R_eff_ for each community as a maximum likelihood estimator for R_eff_ given the past eight days of case reports, assuming Poisson distributed case numbers and exponential growth (accounting for the serial interval mean and standard deviation). Note that this is not intended to be a rigorous inference of the effective reproduction number, but rather a quick proxy that can be used for setting a reasonable threshold. We leave construction of optimal thresholds for local interventions to future work.

4 excluding the Isles of Scilly

5 Lower Layer Super Output Areas

6 School locations were not available for Wales at the time of submission.

7 Primary and secondary schools are treated separately.

8 This is partly due to the fact that MSOA communities are geographically smaller in dense urban environments, leading to greater mixing between communities, and workers and school children are more likely to commute out of their (small) home community for work or school.

9 assuming 64% of contacts traced and isolated on symptom onset.

## References

1. Zhu, N. et al. China novel coronavirus investigating and research team. a novel coronavirus from patients with pneumonia in china, 2019. N Engl JMed 382, 727–733 (2020).

2. Lu, R. et al. Genomic characterisation and epidemiology of 2019 novel coronavirus: implications for virus origins and receptor binding. The Lancet 395, 565–574 (2020).

3. Cai, J. et al. Roles of different transport modes in the spatial spread of the 2009 influenza a (h1n1) pandemic in mainland china. Int. journal environmental research public health 16, 222 (2019).

4. Wang, C., Horby, P. W., Hayden, F. G. & Gao, G. F. A novel coronavirus outbreak of global health concern. The Lancet 395, 470–473 (2020).

5. Flaxman, S. et al. Report 13: Estimating the number of infections and the impact of non-pharmaceutical interventions on covid-19 in 11 european countries. (2020).

6. Tian, H. et al. An investigation of transmission control measures during the first 50 days of the covid-19 epidemic in china. Science (2020).

7. Glasser, J. W., Hupert, N., McCauley, M. M. & Hatchett, R. Modeling and public health emergency responses: Lessons from sars. Epidemics 3, 32–37 (2011).

8. Kang, M. et al. Contact tracing for imported case of middle east respiratory syndrome, china, 2015. Emerg. infectious diseases 22, 1644 (2016).

9. England, P. H. MERS-CoV Close Contact Algorithm. https://assets.publishing.service.gov.uk/government/uploads/system/uploads/attachment_data/file/776218/MERS-CoV_Close_contacts_algorithm.pdf (2019). [Online; accessed 23-April-2019].

10. Swanson, K. C. et al. Contact tracing performance during the ebola epidemic in liberia, 2014–2015. PLoS neglected tropical diseases 12, e0006762 (2018).

11. WHO. Implementation and management of contact tracing for Ebola virus disease. https://www.who.int/csr/resources/publications/ebola/contact-tracing/en/ (2015). [Online; accessed 23-April-2019].

12. Hoang, T. T. T. et al. Active contact tracing beyond the household in multidrug resistant tuberculosis in vietnam: a cohort study. BMC public health 19, 241 (2019).

13. Guidance, E. Risk assessment guidelines for diseases transmitted on aircraft..

14. Ferretti, L. et al. Quantifying sars-cov-2 transmission suggests epidemic control with digital contact tracing. Science (2020).

15. Fraser, C., Riley, S., Anderson, R. M. & Ferguson, N. M. Factors that make an infectious disease outbreak controllable. Proc. Natl. Acad. Sci. 101, 6146–6151 (2004).

16. Peak, C. M., Childs, L. M., Grad, Y. H. & Buckee, C. O. Comparing nonpharmaceutical interventions for containing emerging epidemics. Proc. Natl. Acad. Sci. 114, 4023–4028 (2017).

17. Hellewell, J. et al. Feasibility of controlling covid-19 outbreaks by isolation of cases and contacts. The Lancet Glob. Heal. (2020).

18. Keeling, M. J., Hollingsworth, T. D. & Read, J. M. The efficacy of contact tracing for the containment of the 2019 novel coronavirus (covid-19). *medRxiv* (2020).

19. Li, R. et al. Substantial undocumented infection facilitates the rapid dissemination of novel coronavirus (sars-cov2). Science (2020).

20. Zhang, J. et al. Age profile of susceptibility, mixing, and social distancing shape the dynamics of the novel coronavirus disease 2019 outbreak in china. *medRxiv* (2020).

21. Tindale, L. et al. Transmission interval estimates suggest pre-symptomatic spread of covid-19. *MedRxiv* (2020).

22. Ling, A. & Leo, Y. Potential presymptomatic transmission of sars-cov-2, zhejiang province, china, 2020. (2020).

23. Bai, Y. et al. Presumed asymptomatic carrier transmission of covid-19. Jama (2020).

24. Mizumoto, K., Kagaya, K., Zarebski, A. & Chowell, G. Estimating the asymptomatic ratio of 2019 novel coronavirus onboard the princess cruises ship, 2020. *medRxiv* (2020).

25. Ofcom. Communications Market Report 2019. https://www.ofcom.org.uk/__data/assets/pdf_file/0028/155278/communications-market-report-2019.pdf (2019). [Online; accessed 23-April-2019].

26. Ferguson, N. et al. Report 9: Impact of non-pharmaceutical interventions (npis) to reduce covid19 mortality and healthcare demand. (2020).

27. Siegenfeld, A. F., & Bar-Yam, Y. Eliminating covid-19: A community-based analysis. *arXiv preprint arXiv:2003.10086* (2020).

28. Fang, Y. et al. Sensitivity of chest ct for covid-19: comparison to rt-pcr. Radiology 200432 (2020).

29. Bruning, A. H. et al. Rapid tests for influenza, respiratory syncytial virus, and other respiratory viruses: a systematic review and meta-analysis. Clin. Infect. Dis. 65, 1026–1032 (2017).

30. Lloyd-Smith, J. O., Schreiber, S. J., Kopp, P. E. & Getz, W. M. Superspreading and the effect of individual variation on disease emergence. Nature 438, 355–359 (2005).

31. Mossong, J. et al. Social contacts and mixing patterns relevant to the spread of infectious diseases. PLoS medicine 5 (2008).

32. Xia, Y., Bjørnstad, O. N. & Grenfell, B. T. Measles metapopulation dynamics: a gravity model for epidemiological coupling and dynamics. The Am. Nat. 164, 267–281 (2004).

33. Viboud, C. et al. Synchrony, waves, and spatial hierarchies in the spread of influenza. science 312, 447–451 (2006).

34. Truscott, J. & Ferguson, N. M. Evaluating the adequacy of gravity models as a description of human mobility for epidemic modelling. PLoS computational biology 8 (2012).

35. Davies, N. G. et al. Age-dependent effects in the transmission and control of covid-19 epidemics. *medRxiv* (2020).

36. Backer, J. A., Klinkenberg, D. & Wallinga, J. The incubation period of 2019-ncov infections among travellers from wuhan, china. (2020).

37. Donnelly, C. A. et al. Epidemiological determinants of spread of causal agent of severe acute respiratory syndrome in hong kong. The Lancet 361, 1761–1766(2003).

38. Liu, Y., Funk, S., Flasche, S. et al. The contribution of pre-symptomatic infection to the transmission dynamics of covid-2019. Wellcome Open Res. 5, 58 (2020).

39. He, X. et al. Temporal dynamics in viral shedding and transmissibility of covid-19. *medRxiv* (2020).

40. Li, Q. et al. Early transmission dynamics in wuhan, china, of novel coronavirus–infected pneumonia. New Engl. J. Medicine (2020).

41. Bi, Q. et al. Epidemiology and transmission of covid-19 in shenzhen china: Analysis of 391 cases and 1,286 of their close contacts. *MedRxiv* (2020).

42. Nishiura, H., Linton, N. M. & Akhmetzhanov, A. R. Serial interval of novel coronavirus (covid-19) infections. Int. journal infectious diseases (2020).

43. Du, Z. et al. The serial interval of covid-19 from publicly reported confirmed cases. *medRxiv* (2020).

44. Verity, R. et al. Estimates of the severity of coronavirus disease 2019: a model-based analysis. The Lancet infectious diseases (2020).

45. Buckee, C. O. et al. Aggregated mobility data could help fight covid-19. Sci. (New York, NY) (2020).

46. Tang, M. et al. Contacts in context: large-scale setting-specific social mixing matrices from the bbc pandemic project. https://www.medrxiv.org/content/10.1101/2020.02.16.20023754v2 (2020).

